# The usefulness of D-dimer as a predictive marker for mortality in patients with COVID-19 hospitalized during the first wave in Italy

**DOI:** 10.1101/2022.02.04.22270433

**Authors:** Shermarke Hassan, Barbara Ferrari, Raffaella Rossio, Vincenzo la Mura, Andrea Artoni, Roberta Gualtierotti, Ida Martinelli, Alessandro Nobili, Alessandra Bandera, Andrea Gori, Francesco Blasi, Valter Monzani, Giorgio Costantino, Sergio Harari, Frits R. Rosendaal, Flora Peyvandi, the COVID-19 Network working group

**Author notes:** **Correspondence address** Dr. F. Peyvandi, University of Milan, Department of Pathophysiology and Transplantation, Via Francesco Sforza 35, 20122, Milan, Italy.

## Abstract

**Background:** The coronavirus disease 2019 (COVID-19) presents an urgent threat to global health. Identification of predictors of poor outcomes will assist medical staff in treatment and allocating limited healthcare resources.

**Aims:** The primary aim was to study the value of D-dimer as a predictive marker for in-hospital mortality.

**Methods:** This was a cohort study. The study population consisted of hospitalized patients (age >18 years), who were diagnosed with COVID-19 based on real-time PCR at 9 hospitals during the first COVID-19 wave in Lombardy, Italy (Feb-May 2020). The primary endpoint was in-hospital mortality. Information was obtained from patient records. Statistical analyses were performed using a Fine-Gray competing risk survival model. Model discrimination was assessed using Harrell’s C-index and model calibration was assessed using a calibration plot.

**Results:** Out of 1049 patients, 501 patients had evaluable data. Of these 501 patients, 96 died. The cumulative incidence of in-hospital mortality within 30 days was 20% (95CI: 16%-23%), and the majority of deaths occurred within the first 10 days. A prediction model containing D-dimer as the only predictor had a C-index of 0.66 (95%CI: 0.61-0.71). Overall calibration of the model was very poor. The addition of D-dimer to a model containing age, sex and co-morbidities as predictors did not lead to any meaningful improvement in either the C-index or the calibration plot.

**Conclusion:** The predictive value of D-dimer alone was moderate, and the addition of D-dimer to a simple model containing basic clinical characteristics did not lead to any improvement in model performance.

## Introduction

The coronavirus disease 2019 (COVID-19) is an urgent threat to global health that has severely strained the healthcare system of many countries. [1–3]. Since the outbreak in early December 2019, the number of patients confirmed to have the disease has exceeded 164,981,323 and the number of people infected is probably much higher. More than 3,419,700 people have died from COVID-19 infection (up to May 20^th^ 2021).[4]

Due to a large number of COVID-19 patients overwhelming the Italian healthcare system during the first COVID-19 wave in Lombardy, Italy (Feb-May 2020), it was important to understand the role of early predictive markers, in order to better triage patients.

D-dimer is a fibrin degradation product, which originates from the formation and lysis of cross-linked fibrin and reflects activation of coagulation and fibrinolysis. Among the clinical and biochemical parameters associated with poor prognosis, increased D-dimer levels seemed to be predictive for acute respiratory distress syndrome (ARDS), the need for admission to an intensive care unit (ICU) or death. [5,6] Furthermore, several studies have reported an increased incidence of thromboembolic events in hospitalized COVID-19 patients.[7]

Taken together, these early studies indicate that D-dimer values at admission might be used to determine which patients would require hospitalization. (thereby decreasing the burden on the healthcare system)

Therefore, the primary aim of this paper was to study the predictive value of D-dimer levels at admission on in-hospital mortality. The secondary aim of this paper was assess if there was any causal relationship between D-dimer levels and in-hospital mortality.

## Methods

### Study design and population

This was an observational cohort study. The study population consisted of patients aged > 18 years who were hospitalized and who were positive for COVID-19 based on real-time PCR at 9 Italian hospitals, during the first COVID-19 wave in Lombardy, Italy (Feb-May 2020). Patients that were directly admitted to the ICU were excluded. Patients in this study were followed-up for 30 days.

This study was approved by the Medical Ethics Committee of the Fondazione IRCCS Ca’ Granda Ospedale Maggiore Policlinico. Written informed consent was obtained from patients before data collection. In cases where it was not possible to obtain informed consent, due to severe illness or death. Data collection was still performed assuming the patient’s consent.

### Data collection and definition of variables

All information was obtained from electronic patient records, using a standardized case report form. The exposure of interest, D-dimer levels (expressed as ng/mL) was used as either a continuous variable or split into quartiles with the lowest quartile being used as the reference category. The primary endpoint was in-hospital mortality.

The following patient-and treatment characteristics were obtained: age (continuous variable), sex (dichotomous variable; male, female), the use of anticoagulant therapy during the study (dichotomous variable; yes, no) and the number of days between symptom onset and hospital admission (continuous variable). Lastly, information on the number of comorbidities was obtained (continuous variable; based on the list of comorbidities used in the Charlson comorbidity index, with the addition of clinician-defined obesity). The total list of comorbidities was as follows; cardiovascular disease, chronic obstructive pulmonary disease, chronic kidney disease, diabetes mellitus, cancer, liver disease, dementia, connective tissue disease, HIV aids and clinician-defined obesity.

### Statistical analysis, general approach

Descriptive analyses were reported as mean/SD, median/IQR, or as proportions. The cumulative incidence of in-hospital mortality and the relationship between D-dimer levels and in-hospital mortality were assessed using survival analysis methods. Discharge within 30 days with a good prognosis served as a competing outcome, in that it (practically) precludes the occurrence of the main outcome of interest (in-hospital mortality). Therefore, it was decided to model the relationship between D-dimer and in-hospital mortality using the Fine-Gray competing risk survival model, which accounts for the presence of competing events. In the multivariable analyses, we adjusted for age, sex and the number of comorbidities.

For similar reasons, we did not use the Kaplan Meier function to estimate the cumulative incidence of mortality. Instead, we used the cumulative incidence function, which correctly accounts for competing events.

A complete case analysis was performed, meaning that patients with missing values for the exposure, outcome or confounders were removed.

### Statistical analysis, causal relationship between D-dimer levels and in-hospital mortality

The relationship between D-dimer levels and in-hospital mortality was estimated using the aforementioned Fine-Gray competing risk survival model. We adjusted for age, sex and comorbidities, as well as anticoagulant therapy during hospitalization and the time between symptom onset and hospital admission.

### Statistical analysis, The predictive value of D-dimer for in-hospital mortality

Three prediction models were tested; a model containing only D-dimer as a predictor, a model containing age, sex and comorbidities, and a model containing D-dimer, age, sex and comorbidities.

For all three models, model discrimination (i.e. how well can the model discriminate between patients with and without the outcome) and model calibration (i.e. the degree to which predicted mortality and observed mortality are similar) were assessed.

Model discrimination was measured by calculating a modified version of Harrel’s C-index [8], which is a measure of how well the model can discriminate between patients with and without the outcome. In the presence of competing risks, Harrel’s C-index is biased. [9] We calculated a modified version of Harrel’s C-index by setting the follow-up time of patients who experience a competing event to larger than our prediction horizon (which is 30 days), instead of censoring these patients, as was proposed by Wolbers et al. [9]

Model calibration was measured by first dividing the population into ten groups (or deciles), based on their predicted mortality risk. Next, the predicted 30-day mortality for each decile (obtained from the Fine-Gray competing risk survival model) was plotted against the observed 30-day mortality for that decile (obtained from the cumulative incidence function). Furthermore, to examine calibration across the whole range, we also fitted a LOWESS (Locally Weighted Scatterplot Smoothing) line to the data.

### Sample size calculation

A formal sample size calculation for the development of a prediction model was not performed. However, 96 patients died during follow-up (see results section) and the number of predictors used in the prediction models ranged from 1 (for the model containing only D-dimer as a predictor) to 4 (for the model containing D-dimer, age, sex and comorbidities as predictors). Accordingly, the number of events per predictor ranged from 96 to 24, well above the minimum of 10 events per variable needed to accurately estimate the model coefficients.[10] Therefore, we deemed the sample size sufficient for these analyses.

## Results

### Baseline characteristics

Out of 1094 patients, 506 had missing D-dimer levels, 27 had incomplete follow-up data, 13 were excluded due to immediately being admitted to the ICU after admission and 47 patients had missing data on either anticoagulant therapy during hospitalization, or for the time between symptom onset and hospital admission. Finally, 501 patients had evaluable data. Of these, 96 patients died within 30 days after admission. Patients were enrolled between March 6^th^ 2020 to September 20^th^ 2020. Almost all (98%) patients were enrolled before May 31^st^ 2020. (the end of the first COVID-19 wave) D-dimer values were associated with advanced age and the number of comorbidities at admission. (Table 1) From March 6^th^ 2020 to September 20^th^ 2020, the cumulative incidence of in-hospital mortality within 30 days was 20% (95CI: 16%-23%). Mortality was higher in the early phase of the epidemic and slightly decreased over time. (Supplemental table 1) The cumulative incidence of discharge because of a good prognosis within 30 days was 71% (95CI: 67%-75%) (Figure 1), with most patients (75%) who died doing so in the first 10 days. After this period the death rate slowed down, as evidence by the flattening of the survival curve. (Figure 1)

**Table 1:** Baseline characteristics of COVID-19 patients hospitalized in the region of Lombardy, Italy, during the first COVID-19 wave (Feb-May 2020).

|  | ddimer,<br>< 538 ng/mL<br>(N=126) | ddimer,<br>538-957 ng/mL<br>(N=124) | ddimer,<br>957-1764 ng/mL<br>(N=124) | ddimer,<br>> 1764 ng/mL<br>(N=127) | Overall<br>(N=501) |
| --- | --- | --- | --- | --- | --- |
| mean age (SD) | 57.3 (16.5) | 62.1 (14.1) | 67.8 (14.1) | 69.5 (13.6) | 64.2 (15.4) |
| sex |  |  |  |  |  |
| female | 45 (35.7%) | 31 (25.0%) | 47 (37.9%) | 53 (41.7%) | 176 (35.1%) |
| male | 81 (64.3%) | 93 (75.0%) | 77 (62.1%) | 74 (58.3%) | 325 (64.9%) |
| charlson comorbidity<br>index |  |  |  |  |  |
| no comorbidities | 73 (57.9%) | 74 (59.7%) | 68 (54.8%) | 55 (43.3%) | 270 (53.9%) |
| 1 comorbidity | 36 (28.6%) | 25 (20.2%) | 32 (25.8%) | 41 (32.3%) | 134 (26.7%) |
| 2 comorbidities | 14 (11.1%) | 20 (16.1%) | 16 (12.9%) | 21 (16.5%) | 71 (14.2%) |
| 3 or more<br>comorbidities | 3 (2.4%) | 5 (4.0%) | 8 (6.5%) | 10 (7.9%) | 26 (5.2%) |
| anticoagulant therapy<br>during hospitalization |  |  |  |  |  |
| no | 27 (21.4%) | 26 (21.0%) | 18 (14.5%) | 25 (19.7%) | 96 (19.2%) |
| yes | 99 (78.6%) | 98 (79.0%) | 106 (85.5%) | 102 (80.3%) | 405 (80.8%) |
| mean number of days<br>between first symptoms<br>and admission (SD) | 8.9 (6.5) | 10.7 (12.3) | 11.1 (11.8) | 10.7 (10.0) | 10.4 (10.4) |

SD: standard deviation

**Figure 1:**
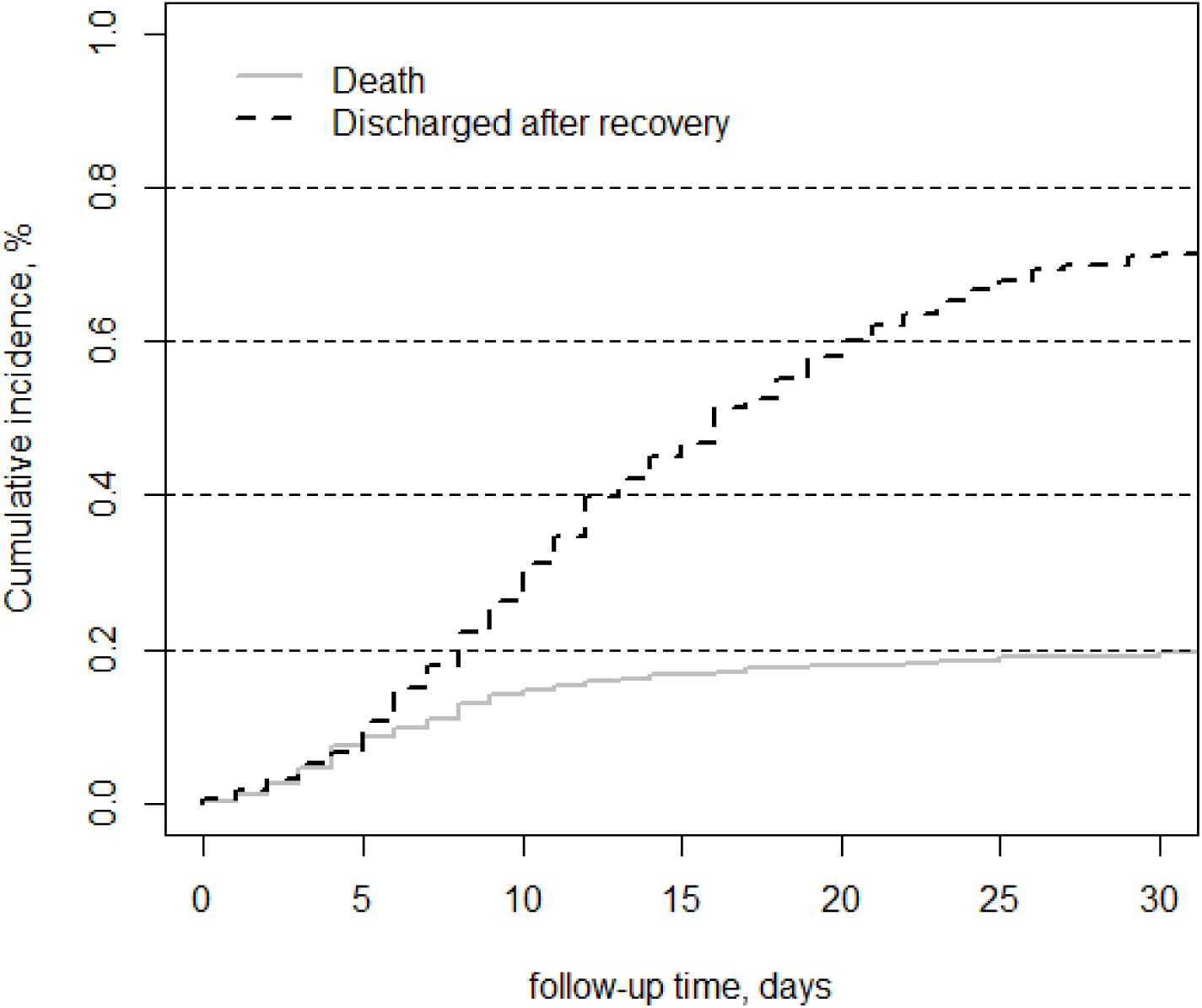
Cumulative incidence function of 501 COVID-19 patients hospitalized in the region of Lombardy, Italy, during the first COVID-19 wave (Feb-May 2020).

With increasing D-dimer levels, the absolute risk of mortality also increased strongly, from 4% (95CI:2%-9%) in patients with D-dimer levels in the lowest quartile to 28% (95CI: 20%-36%) in patients with D-dimer levels in the highest quartile. (Table 2)

**Table 2:**
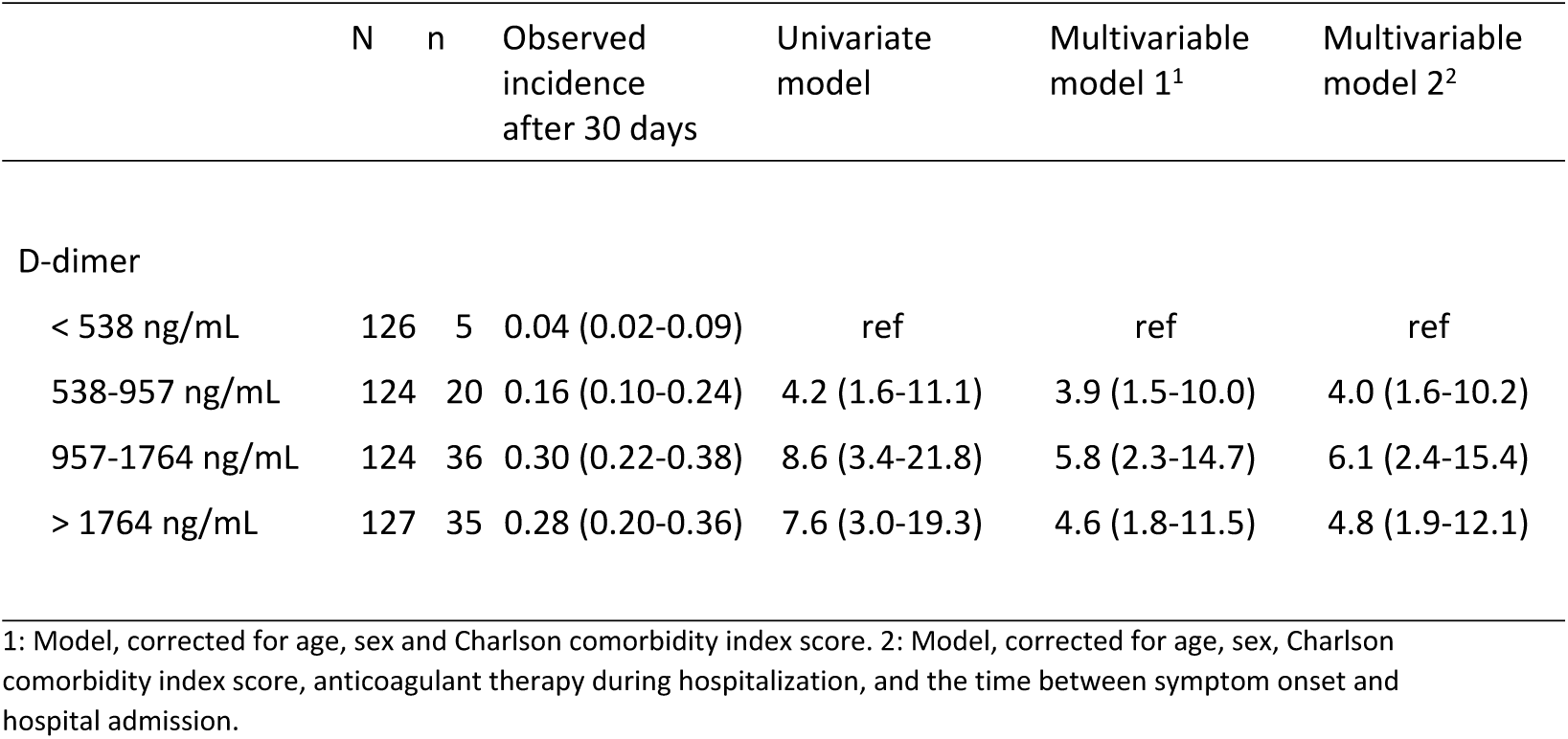
Association between D-dimer values and in-hospital mortality

|  | N | n | Observed<br>incidence<br>after 30 days | Univariate<br>model | Multivariable<br>model 1<sup>1</sup> | Multivariable<br>model 2<sup>2</sup> |
| --- | --- | --- | --- | --- | --- | --- |
| D-dimer |  |  |  |  |  |  |
| < 538 ng/mL | 126 | 5 | 0.04 (0.02-0.09) | ref | ref | ref |
| 538-957 ng/mL | 124 | 20 | 0.16 (0.10-0.24) | 4.2 (1.6-11.1) | 3.9 (1.5-10.0) | 4.0 (1.6-10.2) |
| 957-1764 ng/mL | 124 | 36 | 0.30 (0.22-0.38) | 8.6 (3.4-21.8) | 5.8 (2.3-14.7) | 6.1 (2.4-15.4) |
| > 1764 ng/mL | 127 | 35 | 0.28 (0.20-0.36) | 7.6 (3.0-19.3) | 4.6 (1.8-11.5) | 4.8 (1.9-12.1) |

1: Model, corrected for age, sex and Charlson comorbidity index score. 2: Model, corrected for age, sex, Charlson comorbidity index score, anticoagulant therapy during hospitalization, and the time between symptom onset and hospital admission.

### Causal relationship between D-dimer levels and in-hospital mortality

Compared with patients in the lowest quartile of D-dimer blood concentration, the unadjusted hazard ratio for in-hospital mortality in patients in the 2^nd^, 3^rd^ and 4^th^ quartile was 4.2 (95CI: 1.6-11.1), 8.6 (95CI: 3.4-21.8) and 7.6 (95CI: 3.0-19.3) respectively. (Table 2) After adjusting for age, sex, comorbidities, anticoagulant therapy during hospitalization and the time between symptom onset and hospital admission, the hazard ratio for patients in the 2^nd^, 3^rd^ and 4^th^ quartile was 4.0 (95CI: 1.6-10.2), 6.1 (95CI: 2.4-15.4), and 4.8 (95CI: 1.9-12.1) respectively. (Table 2)

### The predictive value of D-dimer for in-hospital mortality

The predictive model containing D-dimer as the only predictor had a C-index of 0.66 (95%CI: 0.61-0.71). Overall calibration of the model was very poor (Figure 2a). Next, the predictive model containing age, sex and comorbidities as predictors had a C-index of 0.82 (95%CI: 0.78-0.86). Overall calibration of the model was acceptable (Figure 2b). Lastly, the predictive model containing D-dimer, age, sex and comorbidities as predictors had a C-index of 0.83 (95%CI: 0.79-0.86). Overall calibration of this model was acceptable (Figure 2c).

**Figure 2:**
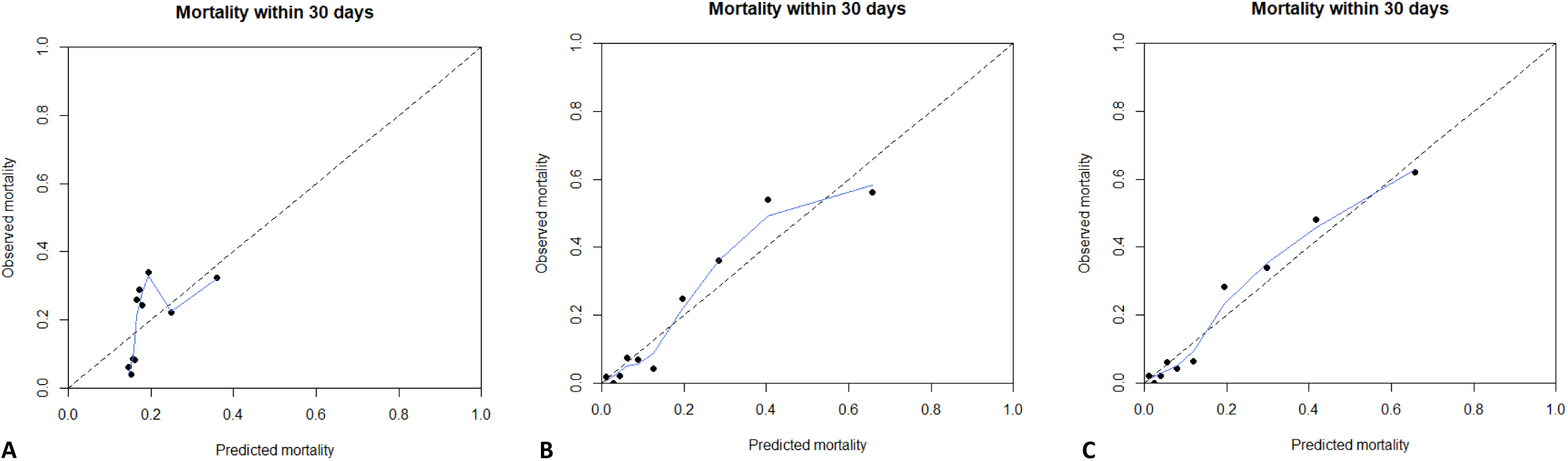
Calibration plot of prediction models *Legend:* The figure shows the calibration plot of the model containing only d-dimer as a predictor (**A**), containing age, sex and comorbidities (**B**) and containing d-dimer, age, sex and comorbidities (**C**). The population was divided into ten groups (or deciles) based on their predicted mortality risk. (represented as black dots in the plot) The predicted probability of mortality according to the model is shown on the X-axis while the observed mortality is shown on the Y-axis. Groups with a higher predicted risk of mortality should have a higher observed risk. To examine calibration across the whole range, we also fitted a LOWESS (Locally Weighted Scatterplot Smoothing) line to the data. (shown here as a blue line) The dotted line represents perfect prediction (where the predicted risk is exactly the same as the observed risk).

## Discussion

Our results show that despite a strong correlation between D-dimer levels and mortality, the predictive value of D-dimer as a single biomarker was unclear. Model discrimination was moderate (C-index: 0.67) while model calibration was very poor. Furthermore, the addition of D-dimer to a simple model containing only basic clinical characteristics (age, sex and co-morbidities) did not lead to any meaningful improvement in either the C-index or the calibration plot.

D-dimer is a breakdown product, generated after a fibrin clot is degraded by fibrinolysis. It is a recognized valid lab biomarker that is widely used as part of the diagnostic workup of patients with a suspected venous thromboembolism (VTE) or disseminated intravascular coagulation (DIC) and is predictive of poor outcomes and thromboembolic events. [11] Changes in D-dimer levels are seen in most patients that are hospitalized with COVID-19. [12] Changes in other hemostatic parameters, such as a slightly elongated PT, elongated aPTT, or mild thrombocytopenia are less common. [13] Furthermore, in addition to the increase in D-dimer (which is also an acute-phase protein that rises with general inflammation) an increase in inflammatory biomarkers such as CRP, particularly in COVID-19 patients with a more severe disease phenotype, is also seen. [14]

The mechanisms underlying this COVID-19 induced coagulopathy may, in part, be explained by the same general mechanisms that also underlie other cases of bacteria-induced septic coagulopathy such as overproduction of pro-inflammatory cytokines by monocytes. Furthermore, direct activation of coagulation by monocytes via tissue-factor and phosphatidylserine (which are expressed on the cell surface of monocytes) also play a role. [14] Furthermore, studies have reported endothelial dysfunction in patients with COVID-19 induced coagulopathy, which is probably mediated by the production of pro-inflammatory cytokines as well as activation of the complement cascade. [15,16]

A strong correlation between D-dimer and mortality was also reported by other studies. A meta-analysis of six studies containing 1355 hospitalized patients found that D-dimer levels were higher in deceased patients (standardized mean difference: 3.59 mcg/L, 95%CI 2.79—4.40). [17] This meta-analysis did not calculate a pooled C-index to assess the overall predictive performance of D-dimer.

A later meta-analysis reporting on 16 studies containing 4468 COVID-19 patients reported a pooled C-index of 0.86 (95CI: 83-89) for predicting all-cause mortality. [18] However, these results were most likely strongly influenced by publication bias. In addition, it is somewhat unclear how the pooled C-index was calculated, as many studies did not report the C-index directly.

A retrospective study by Zhang et al. evaluated D-dimer levels and mortality in 343 patients. [19] D-dimer levels were measured within the first 24 hours, and hospitalized patients were followed until death or discharge. The study showed a very strong correlation between D-dimer levels over 2.0 mcg/mL and mortality. (HR: 51.5, 95%CI 12.9-206.7). However, the study did not adjust for any confounders. It is therefore unclear how different confounders could have affected the reported results. The predictive value of D-dimer was also very high (C-index: 0.89) There was no information about anticoagulant use during the study follow-up.

These strong results were not confirmed by a later study by Naymagon et al. [20] that followed 1062 COVID-19 patients during hospitalization. Each 1 μg/ml increase in D-dimer levels (measured within 3 days of admission) was associated with a hazard ratio of death of 1.05 (95%CI: 1.04-1.07). The association did not change after adjustment for age, smoking, Charlson comorbidity index and anticoagulant use at admission. However, discriminative performance of D-dimer levels was moderate (C-index: 0.694). At baseline, 9.1% of patients were on anticoagulant use and no information was given about anticoagulant use during the study.

Overall, it seems that early studies reported that D-dimer was strongly predictive of mortality, although this effect was not as strong in the larger studies. Furthermore, all aforementioned studies only assessed the discriminative performance of D-dimer, but not model calibration.

Our study has several strengths. Firstly, our study had a large sample size with a sufficient number of events. Secondly, we applied a competing risk survival model to analyze the relationship between D-dimer and poor outcomes to avoid bias. Not taking competing risks into account could lead to misleading results, as shown in a recent simulation study on competing risks in COVID-19 research. [21] In addition, we evaluated both discrimination (as was done in earlier studies) and model calibration (which was not reported in any of the aforementioned studies). This is important because models may show good discrimination but could still be poorly calibrated. [22]

Our study also has some limitations. The main limitation is that values for D-dimer levels were not available for 506 out of 1094 patients. D-dimer tests are most commonly ordered if a patient has some symptoms or medical history which are indicative of a thromboembolic event. Therefore, patients that were excluded from the study due to missing information on D-dimer were most likely patients with a low a priori likelihood of having a VTE. Also, D-dimer assays vary widely in their set-up. This lack of standardization makes comparison of different study results somewhat difficult. [23,24]

Due to the rapid pace of change in the treatment of patients with COVID-19, the predictive value of D-dimer (and therefore, it’s clinical usefulness) will most likely have diminished over time. For example, in Lombardy, many patients in the second COVID-19 wave (Oct-Dec 2020) were already being prescribed anticoagulant treatment by their general practitioner before hospitalization. Furthermore, as the outbreak went on, patients with milder symptoms were also being hospitalized. Due to these treatment changes, we can speculate that patients hospitalized after the first COVID-19 wave will have had lower D-dimer levels at admission, when compared to patients admitted in the first COVID-19 wave (Feb-May 2020). Furthermore, D-dimer levels would have been less strongly associated with mortality in these patients, when compared to patients admitted in the first COVID-19 wave (Feb-May 2020).

As shown before, a part of COVID-19 related mortality is due to an underlying coagulopathy. (which might manifest as a VTE, as DIC or a TMA) Consequently, some studies have suggested that D-dimer levels could be used to stratify patients with COVID-19, and to individualize treatment. [19] However, our analyses show that, despite a strong correlation between D-dimer levels and mortality, the predictive value of D-dimer alone was not sufficient. However, that was to be expected as COVID-19 is not a coagulation disorder but a multi-systemic (although mainly respiratory) disease that influences health through multiple pathways, one of which is the coagulation system. However, D-dimer also showed little added value when added to simple risk prediction model containing only age, sex and comorbidities as predictors.

## Conclusion

The predictive value of D-dimer alone was moderate, and the addition of D-dimer to a simple model containing basic clinical characteristics did not lead to any improvement in model performance.

## Data Availability

For original data, please contact the Angelo Bianchi Bonomi Hemophilia and Thrombosis Centre.

## Data Availability

For original data, please contact the Angelo Bianchi Bonomi Hemophilia and Thrombosis Centre.

## Authorship

### Author contributions

S. Hassan analyzed the data and wrote the manuscript. F. Peyvandi interpreted the data and reviewed the manuscript. F.R. Rosendaal reviewed the manuscript. All other authors were involved in either coordination of the COVID-19 network research project, patient enrollment, data collection and/or reviewing the manuscript.

## Acknowledgments

We thank the COVID-19 Network working group for their help with patient recruitment and data collection. The COVID-19 Network working group consists of the following members:

Silvano Bosari, Luigia Scudeller, Giuliana Fusetti, Laura Rusconi, Silvia Dell’Orto, Daniele Prati, Luca Valenti, Silvia Giovannelli, Maria Manunta, Giuseppe Lamorte, Francesca Ferarri, Andrea Gori, Alessandra Bandera, Antonio Muscatello, Davide Mangioni, Laura Alagna, Giorgio Bozzi, Andrea Lombardi, Riccardo Ungaro, Giuseppe Ancona, Gianluca Zuglian, Matteo Bolis, Nathalie Iannotti, Serena Ludovisi, Agnese Comelli, Giulia Renisi, Simona Biscarini, Valeria Castelli, Emanuele Palomba, Marco Fava, Valeria Fortina, Carlo Alberto Peri, Paola Saltini, Giulia Viero, Teresa Itri, Valentina Ferroni,Valeria Pastore,Roberta Massafra,Arianna Liparoti,Toussaint Muheberimana, Alessandro Giommi, Rosaria Bianco, Rafaela Montalvao De Azevedo, Grazia Eliana Chitani, Flora Peyvandi, Roberta Gualtierotti, Barbara Ferrari, Raffaella Rossio, Nadia Boasi, Erica Pagliaro, Costanza Massimo, Michele De Caro, Andrea Giachi, Nicola Montano, Barbara Vigone, Chiara Bellocchi, Angelica Carandina, Elisa Fiorelli, Valerie Melli, Eleonora Tobaldini, Francesco Blasi, Stefano Aliberti, Maura Spotti,Leonardo Terranova, Sofia Misuraca, Alice D’Adda, Silvia Della Fiore, Marta Di Pasquale, Marco Mantero Martina Contarini, Margherita Ori, Letizia Morlacchi, Valeria Rossetti, Andrea Gramegna, Maria Pappalettera, Mirta Cavallini, Agata Buscemi, Marco Vicenzi, Irena Rota, Giorgio Costantino, Monica Solbiati, Ludovico Furlan, Marta Mancarella, Giulia Colombo, Giorgio Colombo, Alice Fanin, Mariele Passarella, Valter Monzani, Ciro Canetta, Angelo Rovellini, Laura Barbetta, Filippo Billi, Christian Folli, Silvia Accordino, Diletta Maira, Cinzia Maria Hu, Irene Motta, Natalia Scaramellini, Anna Ludovica Fracanzani, Rosa Lombardi, Annalisa Cespiati, Matteo Cesari,Tiziano Lucchi,Marco Proietti, Laura Calcaterra, Clara Mandelli, Carlotta Coppola, Arturo Cerizza, Antonio Maria Pesenti, Giacomo Grasselli, Alessandro Galazzi, Alessandro Nobili, Mauro Tettamanti, Igor Monti, Alessia Antonella Galbussera, Ernesto Crisafulli, Domenico Girelli, Alessio Maroccia, Daniele Gabbiani, Fabiana Busti, Alice Vianello, Marta Biondan, Filippo Sartori, Paola Faverio, Alberto Pesci, Stefano Zucchetti, Paolo Bonfanti, Marianna Rossi, Ilaria Beretta, Anna Spolti, Sergio Harari, Davide Elia, Roberto Cassandro, Antonella Caminati, Francesco Cipollone, Maria Teresa Guagnano, Damiano D’Ardes, Ilaria Rossi, Francesca Vezzani, Antonio Spanevello, Francesca Cherubino, Dina Visca, Marco Contoli, Alberto Papi, Luca Morandi, Nicholas Battistini, Guido Luigi Moreo, Pasqualina Iannuzzi, Daniele Fumagalli and Sara Leone.

## Appendix

**Supplemental Table 1:** cumulative incidence of death, per time-period

| Time period | N | Cumulative incidence of death |
| --- | --- | --- |
| March 6 to September 20 | 501 | 20% (95CI: 16-23) |
| March 6 to March 14 | 35 | 19% (95CI: 7-34) |
| March 14 to March 31 | 263 | 20% (95CI: 15-25) |
| April 1 to April 14 | 126 | 21% (95CI: 14-28) |
| April 14 to April 30 | 51 | 18% (95CI: 9-30) |
| April 30 to September 20 | 26 | 15% (95CI: 5-32) |

## References

1 Arabi YM, Murthy S, Webb S. COVID-19: a novel coronavirus and a novel challenge for critical care. Intensive Care Med 2020; 46: 833–6.

2 Grasselli G, Pesenti A, Cecconi M. Critical Care Utilization for the COVID-19 Outbreak in Lombardy, Italy: Early Experience and Forecast During an Emergency Response. Jama 2020;.

3 Xie J, Tong Z, Guan X, Du B, Qiu H, Slutsky AS. Critical care crisis and some recommendations during the COVID-19 epidemic in China. Intensive Care Med 2020; 46: 837–40.

4 Dong E, Du H, Gardner L. An interactive web-based dashboard to track COVID-19 in real time. Lancet Infect Dis 2020; 20: 533–4.

5 Wu Z, McGoogan JM. Characteristics of and Important Lessons From the Coronavirus Disease 2019 (COVID-19) Outbreak in China: Summary of a Report of 72 314 Cases From the Chinese Center for Disease Control and Prevention. Jama 2020;.

6 Tang N, Li D, Wang X, Sun Z. Abnormal coagulation parameters are associated with poor prognosis in patients with novel coronavirus pneumonia. J Thromb Haemost 2020; 18: 844–7.

7 Al-Ani F, Chehade S, Lazo-Langner A. Thrombosis risk associated with COVID-19 infection. A scoping review. Thromb Res Elsevier Ltd; 2020; 192: 152–60.

8 Harrell FE, Lee KL, Califf RM, Pryor DB, Rosati RA. Regression modelling strategies for improved prognostic prediction. Stat Med Stat Med; 1984; 3: 143–52.

9 Wolbers M, Koller MT, Witteman JCM, Steyerberg EW. Prognostic models with competing risks methods and application to coronary risk prediction. Epidemiology Lippincott Williams and Wilkins; 2009; 20: 555–61.

10 Peduzzi P, Concato J, Kemper E, Holford TR, Feinstein AR. A simulation study of the number of events per variable in logistic regression analysis. J Clin Epidemiol 1996; 49: 1373–9.

11 Weitz JI, Fredenburgh JC, Eikelboom JW. A Test in Context: D-Dimer. Journal of the American College of Cardiology. Elsevier USA; 2017. p. 2411–20.

12 Panigada M, Bottino N, Tagliabue P, Grasselli G, Novembrino C, Chantarangkul V, Pesenti A, Peyvandi F, Tripodi A. Hypercoagulability of COVID-19 patients in intensive care unit: A report of thromboelastography findings and other parameters of hemostasis. J Thromb Haemost Blackwell Publishing Ltd; 2020; 18: 1738–42.

13 Peyvandi F, Artoni A, Novembrino C, Aliberti S, Panigada M, Boscarino M, Gualtierotti R, Rossi F, Palla R, Martinelli I, Grasselli G, Blasi F, Tripodi A. Hemostatic alterations in COVID-19. (Haematologica); 2020; 105: haematol.2020.262634.

14 Iba T, Levy JH, Levi M, Thachil J. Coagulopathy in COVID-19. Journal of Thrombosis and Haemostasis. Blackwell Publishing Ltd; 2020. p. 2103–9.

15 Cugno M, Meroni PL, Gualtierotti R, Griffini S, Grovetti E, Torri A, Lonati P, Grossi C, Borghi MO, Novembrino C, Boscolo M, Uceda Renteria SC, Valenti L, Lamorte G, Manunta M, Prati D, Pesenti A, Blasi F, Costantino G, Gori A, et al. Complement activation and endothelial perturbation parallel COVID-19 severity and activity. J Autoimmun Academic Press; 2021; 116: 102560.

16 Cugno M, Meroni PL, Gualtierotti R, Griffini S, Grovetti E, Torri A, Panigada M, Aliberti S, Blasi F, Tedesco F, Peyvandi F. Complement activation in patients with COVID-19: A novel therapeutic target. J Allergy Clin Immunol Mosby Inc.; 2020; 146: 215–7.

17 Sakka M, Connors JM, Hékimian G, Martin-Toutain I, Crichi B, Colmegna I, Bonnefont-Rousselot D, Farge D, Frere C. Association between D-Dimer levels and mortality in patients with coronavirus disease 2019 (COVID-19): a systematic review and pooled analysis. JMV-Journal de Medecine Vasculaire. Elsevier Masson SAS; 2020. p. 268–74.

18 Zhan H, Chen H, Liu C, Cheng L, Yan S, Li H, Li Y. Diagnostic Value of D-Dimer in COVID-19: A Meta-Analysis and Meta-Regression. Clin Appl Thromb SAGE Publications Inc.; 2021; 27.

19 Zhang L, Yan X, Fan Q, Liu H, Liu X, Liu Z, Zhang Z. D-dimer levels on admission to predict in-hospital mortality in patients with Covid-19. J Thromb Haemost Blackwell Publishing Ltd; 2020; 18: 1324–9.

20 Naymagon L, Zubizarreta N, Feld J, van Gerwen M, Alsen M, Thibaud S, Kessler A, Venugopal S, Makki I, Qin Q, Dharmapuri S, Jun T, Bhalla S, Berwick S, Christian K, Mascarenhas J, Dembitzer F, Moshier E, Tremblay D. Admission D-dimer levels, D-dimer trends, and outcomes in COVID-19. Thromb Res Elsevier Ltd; 2020; 196: 99–105.

21 Oulhaj A, Ahmed LA, Prattes J, Suliman A, Alsuwaidi AR, Al-Rifai RH, Sourij H, Keilegom I Van. The competing risk between in-hospital mortality and recovery: A pitfall in COVID-19 survival analysis research. medRxiv Cold Spring Harbor Laboratory Press; 2020; : 2020.07.11.20151472.

22 Van Calster B, McLernon DJ, Van Smeden M, Wynants L, Steyerberg EW, Bossuyt P, Collins GS, MacAskill P, McLernon DJ, Moons KGM, Steyerberg EW, Van Calster B, Van Smeden M, Vickers AJ. Calibration: The Achilles heel of predictive analytics. BMC Med BioMed Central Ltd.; 2019; 17.

23 Thachil J, Longstaff C, Favaloro EJ, Lippi G, Urano T, Kim PY. The need for accurate D-dimer reporting in COVID-19: Communication from the ISTH SSC on fibrinolysis. J Thromb Haemost Blackwell Publishing Ltd; 2020; 18: 2408–11.

24 Linkins LA, Takach Lapner S. Review of D-dimer testing: Good, Bad, and Ugly. International Journal of Laboratory Hematology. Blackwell Publishing Ltd; 2017. p. 98–103.

